# Remodelling of cystic fibrosis respiratory microbiota in response to extended Elexacaftor–Tezacaftor–Ivacaftor therapy

**DOI:** 10.1101/2025.11.27.25341148

**Authors:** Helen Gavillet, Lauren R Hatfield, Michelle Hardman, Ryan Marsh, Gisli G Einarsson, Christina S Thornton, Michael D Parkins, Jamie Duckers, Jennifer M Bomberger, Yasmin Hilliam, Stella E Lee, Robert W Lord, Andrew Jones, Alexander Horsley, Thomas W V Daniels, Charlotte C Teneback, Damian W Rivett, Christopher van der Gast

**Affiliations:** Department of Applied Sciences, Northumbria University, Newcastle, UK; Department of Life Sciences, Manchester Metropolitan University, Manchester, UK; School of Biological Sciences, University of Manchester, Manchester, UK; School of Pharmacy Queen’s University Belfast, UK; Department of Medicine, Cumming School of Medicine, University of Calgary and Alberta Health Services, Calgary, Canada; Department of Respiratory Medicine, University Hospital Wales, Cardiff and Vale University Health Board, Cardiff, UK; Department of Microbiology and Immunology, Geisel School of Medicine, Dartmouth College, Hanover, New Hampshire, USA; Division of Otolaryngology, Brigham and Women’s Hospital, Boston, Massachusetts, USA; Manchester Adult Cystic Fibrosis Centre, Manchester University NHS Foundation Trust, Manchester, UK; NIHR Respiratory Biomedical Research Centre, Division of Infection, Immunity and Respiratory Medicine, University of Manchester, Manchester, UK; Cystic Fibrosis Unit, Southampton University Hospitals NHS Trust, Southampton, UK; National Institute for Health and Care Research, Southampton Biomedical Research Centre, Southampton UK; Division of Pulmonary and Critical Care Medicine, Department of Medicine, Larner College of Medicine, University of Vermont, USA; Department of Natural Sciences, Manchester Metropolitan University, Manchester, UK; Department of Respiratory Medicine, Northern Care Alliance NHS Foundation Trust, Salford, UK

**Keywords:** Cystic fibrosis, Lung microbiome, Microbiome ecology, Azithromycin, Kaftrio, Trikafta, CFTR modulator therapy, dysbiosis

## Abstract

**Background:** Cystic fibrosis (CF) has profoundly changed since the introduction of CF Transmembrane Conductance Regulator modulator therapies (CFTRmt), a class of medications that improve function of the CFTR protein encoded by certain CF-causing gene mutations. Amongst these, the triple combination therapy elexacaftor-tezacaftor-ivacaftor (ETI) has been the most impactful and widely used to date. Given chronic respiratory infection and concomitant inflammation is the leading cause of morbidity and early mortality for the majority in CF, what is not certain are the long-term effects of ETI therapy on the respiratory microbiota and pathogens imbedded within. Here we assessed the long-term effects of ETI CFTRmt over 3-years on the respiratory microbiota of a multi-centre cohort of 276 adults with CF (awCF) from 6 CF centres in the UK, USA, and Canada, and compared to a non-CF healthy cohort.

**Results:** We determined that respiratory microbiota characteristics (diversity, dominance, and composition) became decreasingly like those of awCF pre-ETI and remodelled to align more with the healthy cohort, where canonical CF pathogens increasingly became less ecologically important in terms of their distributions and abundances across awCF with increased duration on therapy. However, the on-ETI microbiota was impeded from becoming fully ‘healthy’ due to continued antibiotic exposure and irreversible lung damage experienced by awCF. Specifically, we found that azithromycin, an antibiotic widely used principally for its immunomodulatory benefits, had adverse effects on the respiratory microbiota nullifying the observed positive effects of ETI treatment. When administered alongside ETI-therapy, the use of azithromycin maintained a pre-ETI microbiota dysbiosis and enabled enhanced persistence of emblematic CF pathogens.

**Conclusions:** The highly anticipated introduction of ETI CFTRmt has greatly changed the course of CF for many people living with this inherited disease. Here we find that ETI CFTRmt enabled positive remodelling of the respiratory microbiota towards a healthy-like state. However, azithromycin impeded total remodelling, making it an ideal candidate for evaluation for discontinuation in the CFTRmt era. While traditional pathogens become less ecologically important the potential evolution and emergence of virulent strains should be investigated. Additionally, the impacts and implications of ETI therapy on the understudied fungal microbiota should also be explored.

## Background

Cystic fibrosis (CF) is historically a life-limiting inherited genetic disorder [1]. Mutations in the CF transmembrane conductance regulator (CFTR) gene cause functional defects in the encoded epithelial cell apical membrane anion channel leading to defective chloride ion transport, airway surface liquid depletion, and impaired or absent mucociliary clearance [1, 2]. Although CF is a multi-systemic disorder, the primary cause of morbidity and early mortality relate to a vicious cycle of chronic respiratory infection and concomitant inflammation that gradually damages lung tissue and degrades lung function throughout the life of a person living with CF [3–5]. Respiratory infections in CF are particularly challenging as they do not adhere to traditional ‘one microbe, one disease’ concepts of infection pathogenesis. Instead, CF airway infections are comprised of diverse and dynamic interacting microbiota that are highly personalised to the individual [3–6]. However, despite the highly personalised nature of the lung microbiome, a well-established ecological pattern in CF is one where respiratory microbiota diversity and dominance decrease and increase respectively with reductions in lung function and hence with more severe disease [4, 7]. Accordingly, the microbiota of individual people with CF become progressively dominated by canonical CF pathogens with worsening lung function (from mild to moderate to severe) [4]. As such, in combination with measures of lung function, microbiota diversity and dominance, along with the identity of the dominant bacterial species, have been recognized as informative indicators of disease state in CF [4].

CF has profoundly changed since the introduction of CFTR modulator therapies (CFTRmt), a class of medications that improve transport and function of the CFTR protein encoded by certain CFTR gene mutations [2, 8]. Although various CFTRmt have become available over the last decade, the triple combination therapy elexacaftor-tezacaftor-ivacaftor (ETI) has been the most impactful to date [8]. Improvements in lung function, reduced frequency of acute pulmonary exacerbations, and increased body mass index have all been associated with receiving ETI therapy [8]. ETI is now available in the UK, USA, and Canada for people living with CF aged 2 years and above who have at least one copy of the F508del CFTR mutation [9–11]. Furthermore, F508del is the most common disease-causing mutation in CF, with over 85% of pwCF in the UK and North America carrying at least one copy of F508del. As such, ETI has the widest reach across the CF population compared to previous CFTRmt [9–11].

To date there have been a limited number of studies examining the effects of ETI therapy on the respiratory microbiota and the CF pathogens contained within [12–20]. These studies have been based on relatively small patient cohorts of adolescents (≥ 12 years) or adults with CF (awCF) (mean number of participants ± SD = 44.0 ± 53.9, minimum = 7, maximum = 186), from single centres (except [15]), over relatively short sampling periods on ETI therapy (mean duration ± SD = 302.0 ± 141.5 days, minimum = 88 days, maximum = 580 days) [12–20]. In general, most of those studies found the abundance of *Pseudomonas aeruginosa* was reduced on ETI therapy compared to pre-ETI baseline but was not commonly eradicated [12, 13, 15–17, 20]. Similarly, other CF pathogens were reported to have reduced abundance, including *Staphylococcus aureus* [15, 16], *Burkholderia cepacia* complex [20], and *Stenotrophomonas maltophilia* [15]. Generally, microbiota diversity was found to increase, and microbiota composition significantly changed when on ETI therapy compared to baseline [14–19]. However, only one study compared CF participants (*n* = 79) on ETI with healthy participants (*n* = 10), finding the lung microbiota became more healthy-like in terms of diversity and composition but remained significantly different throughout the 12 month study duration [17]. A majority consensus from those studies was a need to further study and better understand the long-term implications of ETI therapy on the CF respiratory microbiome [21].

In the current study, we assessed the long-term effects of ETI CFTRmt over 3+ years on the respiratory microbiota of a large multi-centre cohort of 276 adults from six CF centres located in the UK, USA, and Canada (Table 1). First, we took all collected samples as independent of each other, then stratified all pre-ETI samples by lung disease severity (severe, moderate, and mild) and stratified all on-ETI samples by duration on therapy (6 months, 1 year, 2 years, and 3 years). We then compared respiratory microbiota characteristics (diversity, dominance, and composition) and CF pathogen distributions amongst the stratified pre-ETI and on-ETI samples, along with respiratory samples from a cohort of non-CF healthy controls. Next, we compared the respiratory microbiota within paired pre-ETI and on-ETI respiratory samples to investigate individual responses to this CFTRmt and concomitant clinical factors. Finally, as not all people with CF produce sputum regularly, for example those with mild CF disease or following the effects of ETI therapy [14, 22], we assessed the influence of sample type (sputum and cough swab) on microbiota characteristics.

**Table 1.**
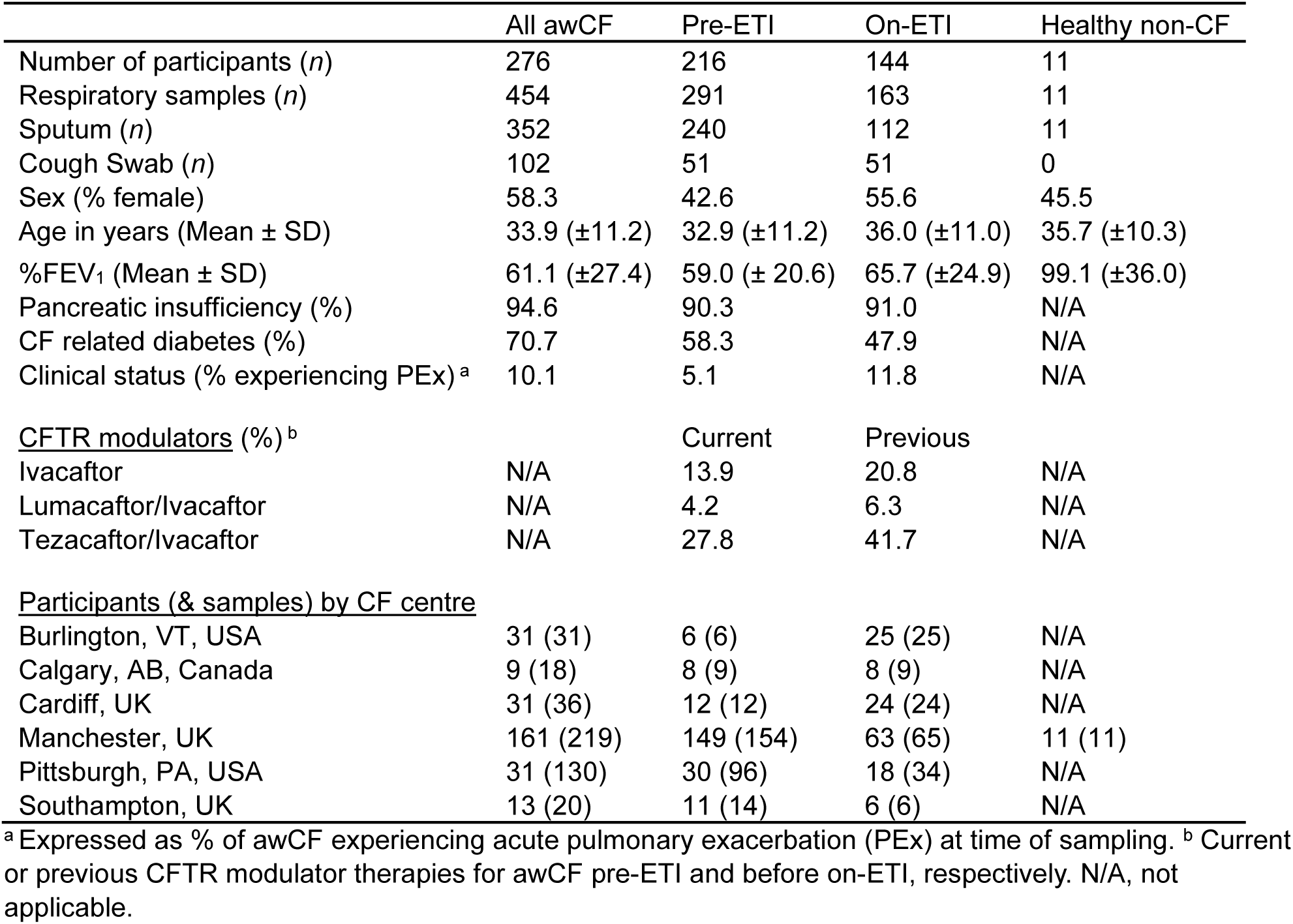
Clinical characteristics for all participants, including adults with CF (awCF) before or on-elexacaftor/tezacaftor/ivacaftor (ETI) therapy and healthy adult controls.

## Methods

### Study and patient sampling

Adults with CF were recruited from six participating centres in the UK, USA, and Canada, with the clinical characteristics of patients included in the study summarised in Table 1. There were no recruitment restrictions other than having approved CFTR mutations for ETI therapy and age (≥ 18 years at start of study). The study was reviewed and approved for each participating centre: Burlington, VT, USA (University of Vermont Institutional Review Board, CHRMS STUDY683), Calgary, AB, Canada (University of Calgary’s Conjoint Health Research Ethics Board, REB-15–2744), Cardiff, UK (Health and Care Research Wales Research Ethics Committee, 18/WA/0089), Manchester, UK (North West – Preston Research Ethics Committee, 20/NW/0302 and NHS Research Ethics Committee North West, Lancaster, 14/NW/1195), Pittsburgh, PA, USA (University of Pittsburgh Institutional Review Board, STUDY19100149), and Southampton, UK (South Central – Hampshire A Research Ethics Committee, 08/H0602/126). All patients attending each participating centre provided written informed consent.

Spontaneously expectorated sputum samples or cough swab samples (if not sputum productive) were collected from participating awCF at their clinic appointments by their regular clinical team and included routine and emergency visits [22]. Induced sputum samples were collected from non-CF healthy participants as previously described [23]. For awCF samples were collected pre-ETI (within 3 months prior to commencement of ETI therapy) and on ETI over a period of approximately 3.5 years duration on therapy (maximum duration 1313 days). Respiratory samples were stored within 3 hours at −80°C prior to DNA extraction [24, 25].

### Sequencing

Nucleic acid extraction was performed on sputum and cough swab samples as previously described, with a modification for the latter sample type [26]. As an alternative to the wash stage for sputum, cough swabs were saturated in sterile phosphate buffer solution for 5 min, then squeezed with sterile tweezers to extract as much material as possible. The resulting solution was then introduced at the bead beating stage and the protocol continued as normal as for the sputum samples thereafter.

Following DNA extraction, approximately 2 ng of template DNA was amplified using Q5 high-fidelity DNA polymerase (New England Biolabs, Hitchin, UK) using a paired-end sequencing approach targeting the V4 bacterial 16S rRNA gene region as previously described [27]. Pooled barcoded amplicon libraries were sequenced on the Illumina MiSeq platform (V3 chemistry). Mock communities, DNA extract, and PCR negative controls were included in each sequencing run [28]. Sequence processing and analysis were carried out in R (Version 4.0.1), utilising the package DADA2, as previously described [29]. Raw sequence data have been deposited in the European Nucleotide Archive under study accession number PRJEB95094. All sequences were putatively assigned genus or species level identification by using the GTDB database [30] and then any remaining non-assigned ASVs were run through BLAST [31]. Given the length of the ribosomal sequences analysed, species identities should be considered putative. A sequence match of 97% or more when run through the databases was required for identification.

### Statistical analysis

Pre-ETI samples were stratified by disease severity based on lung function (percent predicted forced expiratory volume in 1 s (%FEV_1_)) into severe (%FEV_1_ <40%), moderate (%FEV_1_ 40-69%), and mild (%FEV_1_ ≥70%) categories, as previously described [4]. On-ETI samples were stratified by duration on ETI into 6 months (from 0 to 6 months on ETI), 1 year (± 6 months), 2 years (± 6 months), and 3 years (± 6 months and any samples beyond [of which included 2 samples at 1265 and 1313 days on ETI]).

Microbiota diversity was measured using the Shannon (*H*’) and Simpson’s (*D*) indices of diversity. As *H*’ is narrowly constrained in most instances making interpretation difficult we used its exponential form (e*H’*) to overcome that issue [32]. Likewise, *D* is also noted to be constrained and can have variance problems. Therefore, we used the recommended transformation from *D* to -ln(*D*) that also has the additional benefit of being independent of the number of sequence reads per sample [33]. Dominance was measured using the Berger-Parker index of dominance and microbiota compositional similarities measured using the Sørensen index of similarity. All indices were calculated using PAST v4.17 [34]. SIMPER and PERMANOVA analyses were also performed in PAST. All regression analyses, coefficients of determination (*R*^2^), degrees of freedom, *F*-statistics, and significance (*P*) were calculated using XLSTAT v2018.1 (Addinsoft, Paris, France). Kruskal-Wallis analyses in conjunction with the post hoc Dunn tests, were performed in XLSTAT. Direct ordination, by means of canonical correspondence analysis (CCA), was used to relate variability in microbiota composition to clinical and demographic factors. CCA was performed in CANOCO v5 [35]. Clinical/demographic variables that significantly explained variation were determined with forward selection (999 Monte Carlo permutations with false discovery rate) and used in CCA [36].

## Results

Here we analysed respiratory samples from 276 adult CF patients prospectively collected before and on ETI therapy along with samples from 11 non-CF healthy adults. The clinical characteristics of individual patients are summarised in Table 1. A total of 454 samples were collected, of which 102 were cough swabs and 352 were sputum. Microbiota characteristics including indices of diversity, dominance, and compositional similarity were found not to be significantly different by sample type (Kruskal-Wallis (*H*) and significance (*P*): Shannon, *H* = 0.341, *P* =0.559; Simpson’s, *H* = 0.133, *P* = 0.715; Berger-Parker, *H* = 1.115, *P* =0.291; and Sørensen, *H* = 0.271, *P* = 0.603 (Figure S1). However, sequence reads per sample were found to be significantly higher in sputum samples (*H* = 51.81, *P* <0.0001) (Figure S1).

When all 454 patient samples were taken as independent of each other and stratified by disease severity pre-ETI therapy (severe *n* = 65, moderate *n* = 131, and mild *n* = 95) or by duration on ETI therapy (6 months *n* = 46, 1 year *n* = 39, 2 years *n* = 38, and 3 years *n* = 40) diversity and dominance within pre-ETI samples were found to increase and decrease respectively with decreasing severity (Figure 1). Similarly, diversity increased, and dominance decreased with duration on ETI therapy. All diversity and dominance indices were significantly higher or lower, respectively, at years 2 and 3 duration on ETI when compared to pre-ETI samples within the severe, moderate and mild stratifications (*P* < 0.05 in all instances, Figure 1 and Table S1). Moreover, all stratified groups were significantly different to the diversity and dominance observed within the healthy cohort respiratory microbiota, expect for Simpson’s index year 2 on-ETI (*H* = 3.717, *P* = 0.054) and Berger-Parker index for years 2 and 3 (*H* = 0.628, *P* = 0.428 and *H* = 2.795, and *P* = 0.095, respectively). A decrease in dominance of CF pathogens was also observed with decreasing severity in the pre-ETI samples and with increasing duration on-ETI therapy (Figure 1).

**Figure 1.**
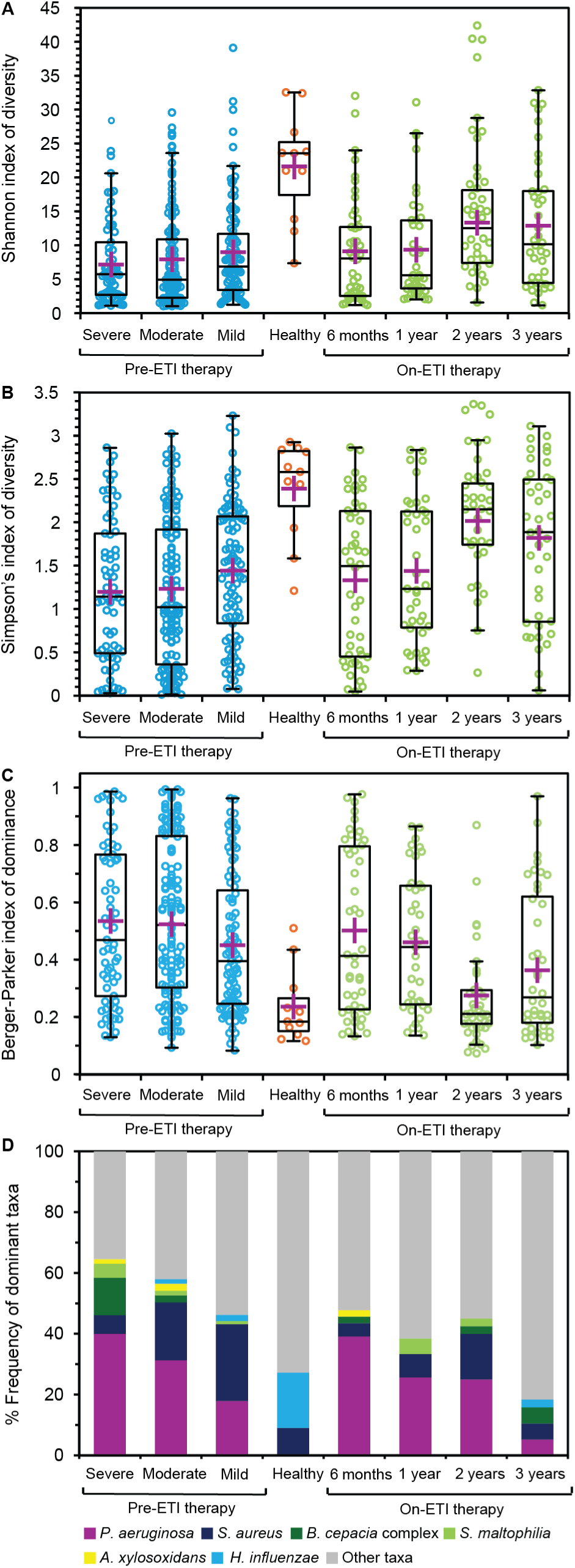
Comparison of respiratory microbiota diversity and dominance pre- and on-Elexacaftor–Tezacaftor–Ivacaftor (ETI) therapy. (A) Shannon index of diversity, expressed as e*^H’^*, Simpson’s index of diversity, expressed as -ln(*D*). (C) Berger-Parker index of dominance (*d*), and (D) Frequency of dominant bacterial taxa in each group (defined as the most abundant taxon by relative abundance within each given microbiota sample). Pre-ETI samples have been stratified by lung disease category, on-ETI samples have been stratified by therapy duration, and a non-CF healthy cohort has been included. Boxplots show 25-75^th^ interquartile (IQR) range with whiskers showing 1.5 times IQR. Purple crosses represent the mean in each group. Circles denote pre-ETI (blue circles), healthy (orange), and on-ETI therapy (green) samples. Summary statistics for Kruskal-Wallis tests between all groups taken pairwise for all indices are provided in Supplementary Table S1.

In terms of microbiota composition, SIMPER analysis revealed which bacterial taxa contributed most between pre-ETI, on-ETI and healthy microbiota dissimilarities (Table 2). The canonical CF pathogens, *P. aeruginosa*, *B. cepacia* complex, *S. maltophilia*, and *Achromobacter xylosoxidans* all followed a trend of lower abundance with improving disease state pre-ETI. Conversely, *S. aureus* and *Haemophilus influenzae* increased in abundance. On-ETI, *P. aeruginosa* and *A. xylosoxidans* decreased in abundance with increasing duration on ETI, with the latter undetectable in any awCF by year 3. *S. maltophilia* also demonstrated a similar trend of abundance reduction with therapy duration. Whereas the *B. cepacia* complex fluctuated in abundance between a maximum of 2.03% and minimum of 0.60%, while both *S. aureus* and *H. influenzae* demonstrated trends of increasing abundance with therapy duration (with the former decreasing in abundance in year 3) (Table 2). Strikingly, all CF pathogens reduced in persistence across awCF with duration on ETI, but by year 3 *H. influenzae* was the only CF pathogen to retain core taxa status (Figure 2 and Table S2). Microbiota composition was observed to be significantly different between pre-ETI and on ETI stratified groups (*P* < 0.05 in all instances) (Figure S2 and Table S3). Further, microbiota composition was observed to become increasingly like the healthy cohort with duration on ETI therapy but remained significantly different (*P* < 0.05) throughout.

**Figure 2.**
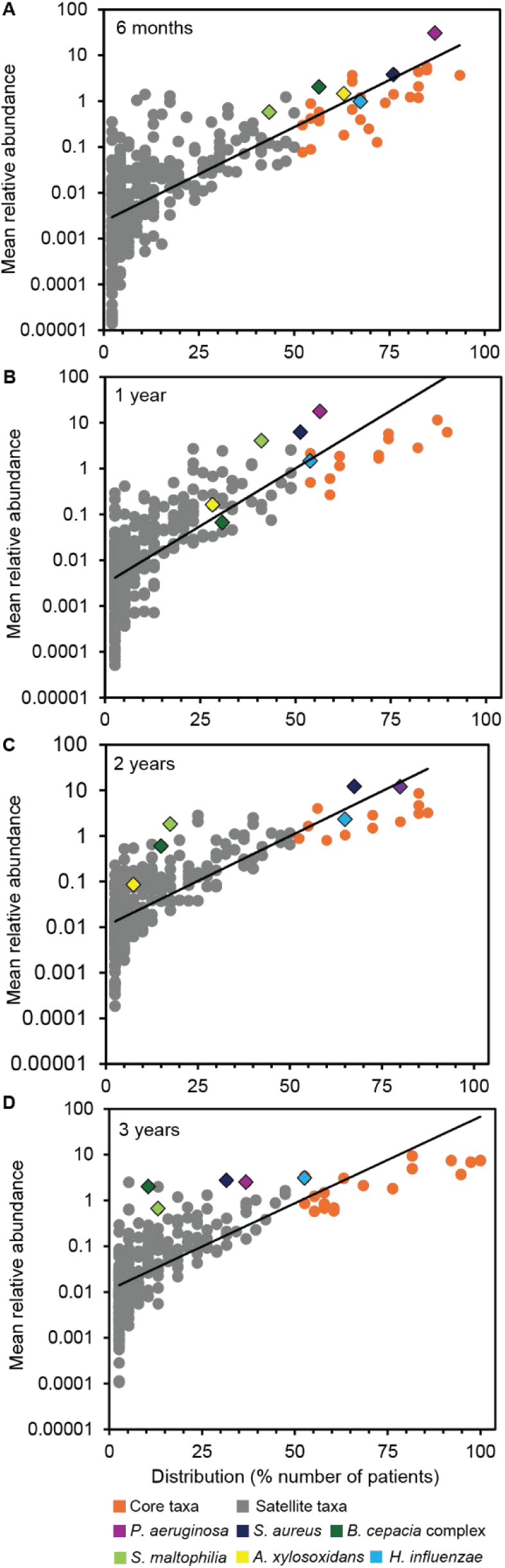
Distribution and abundance of bacterial taxa across adults with CF by duration on-ETI therapy. (**a**) 6 months duration on therapy. (**b**) 1 year. (**c**) 2 years, and (**d**) 3 years. Given is the percentage number of patient respiratory samples each bacterial taxon was observed to be distributed across, plotted against the mean percentage abundance across those samples. Core taxa are defined as those are in >50% of samples (orange circles), and satellite taxa (grey circles) defined as those that do not. Canonical CF pathogens are highlighted in each plot. Distribution-abundance relationship regression statistics: (**a**) *R*^2^ = 0.55, *F*_1,325_ = 395.1, *P* < 0.0001; (**b**) *R*^2^ = 0.57, *F*_1,313_ = 418.1, *P* < 0.0001; (**c**) *R*^2^ = 0.57, *F*_1,268_ = 366.4, *P* < 0.0001; and (**d**) *R*^2^ = 0.55, *F*_1,262_ = 316.1, *P* < 0.0001. Core taxa are listed Supplementary Table S2.

**Table 2.**
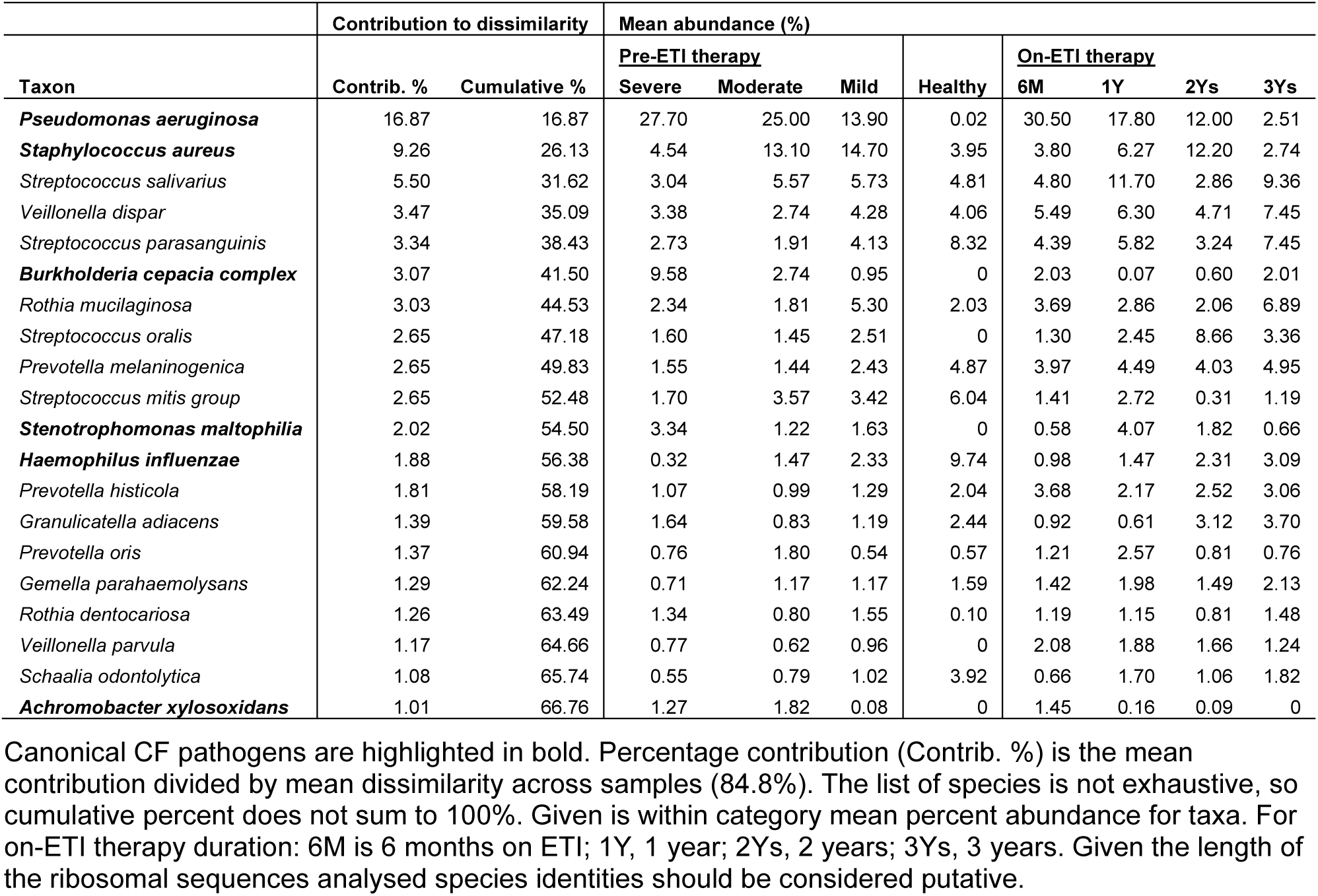
Pooled similarity (SIMPER) analysis of microbiota between pre-ETI, healthy, and on-ETI therapy groups.

To investigate individual responses to ETI CFTRmt and concomitant clinical factors we analysed paired pre-and on-ETI samples from 82 awCF. These awCF were from 5 of the 6 study centres (Calgary *n* = 7; Cardiff *n* = 5; Manchester *n* = 52, Pittsburgh *n* = 13, and Southampton *n* = 5) with pre-ETI samples distributed across disease severity categories (severe *n* = 25, moderate *n* = 38, mild *n* = 19) and across on-ETI duration categories (6 months *n* = 31, 1 year *n* = 20, 2 years *n* = 12, 3 years *n* = 40). Lung function and microbiota diversity and dominance metrics were compared between pre-ETI and on-ETI samples when stratified into pre-ETI disease categories (Figure 3 and Table S4). Lung function was observed to significantly increase on-ETI when comparted to pre-ETI within each pre-ETI disease category (Kruskal Wallis: Severe *H* = 16.28, *P* < 0.0001; moderate *H* = 10.48, *P* < 0.001; and mild *H* = 5.00, *P* = 0.025). Furthermore, the mean percentage change in %FEV_1_ increased with increasing disease severity (severe mean ± SD = 44.2 ± 32.8%, moderate = 14.3 ± 20.9%, and mild = 10.9 ± 8.0%). Generally, diversity increased on-ETI within each pre-ETI disease category. But these differences were only significant in the mild disease category for each diversity index (Shannon *H* = 5.52, *P* = 0.019 and Simpson’s *H* = 4.73, *P* = 0.030). For both diversity indices the mean percentage change increased with decreasing disease severity (Severe Shannon mean ± SD = 31.8 ± 89.3%, moderate = 118.5 ± 260.3%, and mild = 273.9 ± 434.7%; Severe Simpson’s mean ± SD = 191.6 ± 787.3%, moderate = 266.2 ± 520.2%, and mild = 328.0 ± 760.9%). For dominance, although Berger-Parker indices decreased with each disease severity category on-ETI compared to pre-ETI, there was no pattern of change across categories, e.g. from severe to mild as observed for diversity.

**Figure 3.**
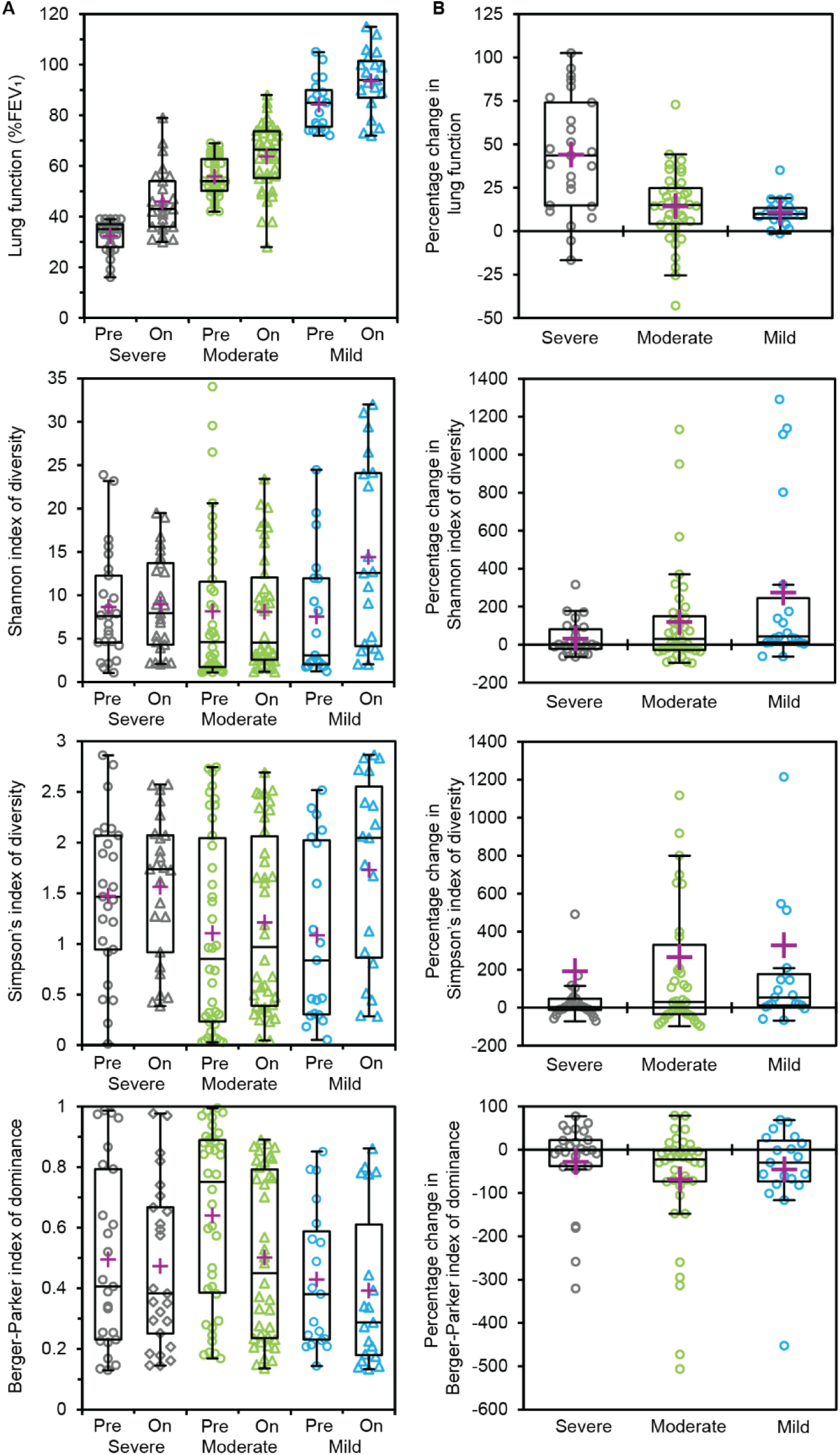
Comparison of lung function and microbiota diversity and dominance metrics between paired pre-and on-ETI therapy samples when stratified by pre-ETI disease severity status. (**A**) Comparison of lung function and microbiota diversity and dominance between pre- and on-ETI samples. (**B**) Percentage change in lung function and microbiota diversity and dominance measures. Circles denote severe (grey circles), moderate (green), and mild (blue) disease severity. Boxplots show 25-75^th^ interquartile (IQR) range with whiskers showing 1.5 times IQR. Purple crosses represent the mean in each group. Summary statistics for Kruskal-Wallis tests are provided in Supplementary Table S4.

To determine which clinical factors significantly accounted for variation in respiratory microbiota composition we performed direct ordination using canonical correspondence analysis for all paired samples, pre-therapy samples only, and on therapy samples only (Table 3). ETI therapy significantly accounted for microbiota composition across all samples, while disease severity and duration on ETI therapy significantly accounted for microbiota variation in the pre-therapy and on-therapy only samples, respectively. Moreover, lung function and antibiotics (azithromycin and tobramycin) significantly accounted for microbiota variation in all instances.

**Table 3.** Canonical correspondence analysis for determination of percent variation in the respiratory microbiota by significant clinical variables between all paired samples, and pre-therapy and on-therapy samples.

|  | All samples |  |  | Pre-therapy |  |  | On-therapy |  |  |
| --- | --- | --- | --- | --- | --- | --- | --- | --- | --- |
|  | Var. exp (%) | pseudo-*F* | *P*(adj) | Var. exp (%) | pseudo-*F* | *P*(adj) | Var. exp (%) | pseudo-*F* | *P*(adj) |
| On/off therapy | 8.0 | 3.2 | 0.002 |  |  |  |  |  |  |
| Disease severity |  |  |  | 5.4 | 1.4 | 0.036 |  |  |  |
| Duration on therapy |  |  |  |  |  |  | 10.5 | 2.8 | 0.004 |
| Lung function | 4.2 | 2.2 | 0.012 | 6.4 | 1.6 | 0.04 | 6.9 | 1.9 | 0.012 |
| CF related diabetes |  |  |  | 1.8 | 1.5 | 0.022 |  |  |  |
| Azithromycin | 5.1 | 2.8 | 0.002 | 5.7 | 1.5 | 0.032 | 8.4 | 2.2 | 0.008 |
| Tobramycin | 6.9 | 3.8 | 0.002 | 5.1 | 1.4 | 0.038 | 9.6 | 2.6 | 0.002 |
| Colistin | 3.9 | 2.1 | 0.004 |  |  |  |  |  |  |
| Total | 28.1 |  |  | 24.4 |  |  | 35.4 |  |  |

Var. exp (%) the percentage of microbiota variation explained by a parameter given in the canonical correspondence analysis. Pseudo-*F* is a test statistic. *P*(adj) adjusted significance value following false discovery rate correction. Non-significant clinical input variables included age, sex, samples type (swab/sputum), originating CF centre, pancreatic insufficiency, clinical status (stable/exacerbation), CFTR genotype, CFTR genotype class, mutation type, and current or previous CFTR modulator for pre-ETI and on-therapy analysis, respectively.

Next changes in microbiota compositional similarity across paired pre- and on-therapy samples were analysed (Figure 4). Microbiota similarity was found to increase with decreasing pre-ETI disease severity (severe mean similarity ± SD = 0.42 ± 0.24, moderate = 0.49 ± 0.23, and mild = 0.53 ± 0.21), while similarity was observed to decrease from pre-ETI compositions with increasing duration on ETI therapy (6 months mean similarity ± SD = 0.62 ± 0.16, 1 year = 0.55 ± 0.19, 2 years = 0.39 ± 0.24, and 3 years = 0.24 ± 0.15). Compositional similarity between paired pre- and on-therapy samples was found to be significantly more similar for awCF receiving azithromycin (*n* = 46) compared to those who were not (*n* = 36) (receiving azithromycin mean similarity ± SD = 0.57 ± 0.36, not receiving = 0.21 ± 0.21)(*H* = 15.48, *P* < 0.0001). All of those receiving azithromycin were distributed across the pre-ETI disease severity categories (severe *n* = 13, moderate *n* = 21, and mild *n* = 12).

**Figure 4.**
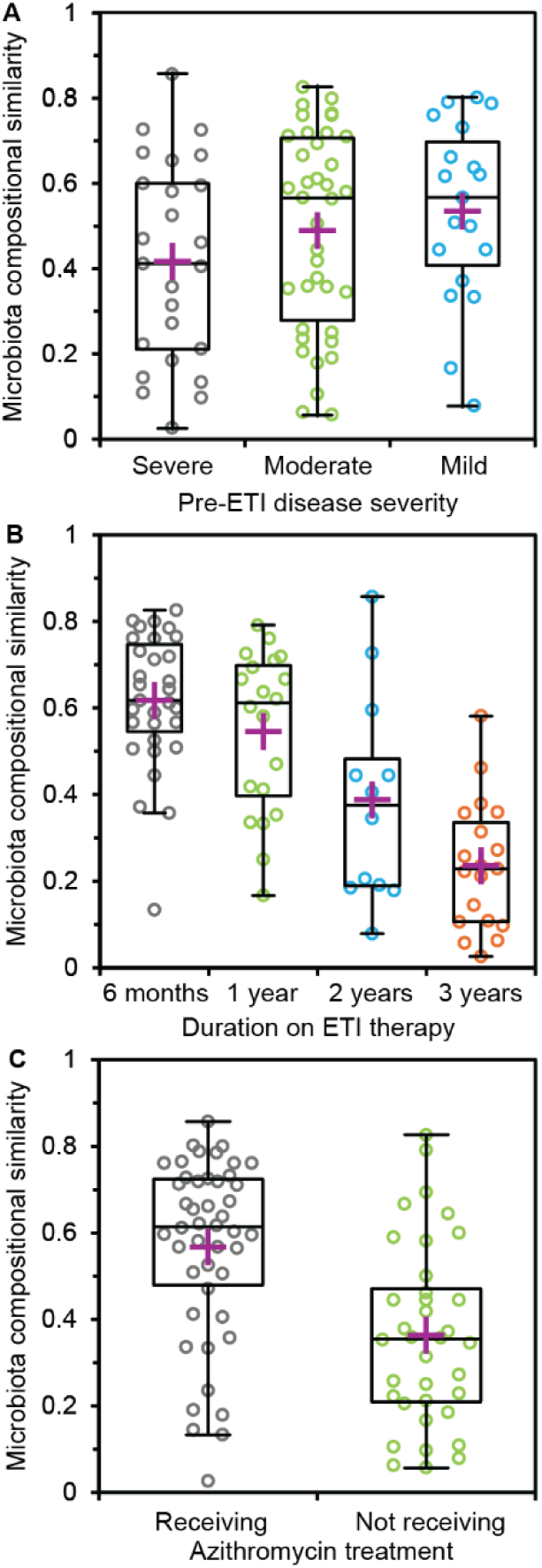
Changes in microbiota composition across paired pre- and on-ETI therapy samples. Given are compositional similarities when stratified by (**A**) pre-ETI disease severity, (**B**) on-ETI therapy duration, and (**C**) azithromycin treatment. Boxplots show 25-75^th^ interquartile (IQR) range with whiskers showing 1.5 times IQR. Purple crosses represent the mean in each group. Compositional similarities were measured using the Sørensen index of similarity.

Azithromycin treatment was also found to significantly influence the distributions and abundances of canonical CF pathogens (Figure 5). The distributions and abundances of all pathogens (except *H. influenzae*) were observed to decrease in awCF with the transition from pre-ETI to on-ETI therapy (mean ± SD reduction in distribution and abundance was 5.6 ± 3.7% and 3.1 ± 3.2%, respectively), with *H. influenzae* distribution and abundance increasing by 9.8% and 0.7%, respectively. Conversely, when azithromycin treatment was accounted for, the distribution of all pathogens was greater across the awCF receiving that systemic antibiotic compared to those that were not for both pre-ETI and on-ETI therapy (mean ± SD increase = 30.1 ± 13.7% and 23.7 ± 9.7%, respectively). For pre-ETI therapy, pathogen abundances were found to increase with azithromycin treatment for *P. aeruginosa* (7.6%), *S. aureus* (4.1%), *B. cepacia* complex (0.6%), and *A. xylosoxidans* (4.3%). Whereas *S. maltophilia* and *H. influenzae* decreased by 4.6% and 3.0%, respectively, in comparison. Similar was observed for on-ETI therapy with increases for *P. aeruginosa* (7.4%), *B. cepacia* complex (1.1%), and *A. xylosoxidans* (1.2%), and *H. influenzae* 2.1% greater), while *S. aureus* and *S. maltophilia* decreased in comparison (-2.0% and -2.6%, respectively).

**Figure 5.**
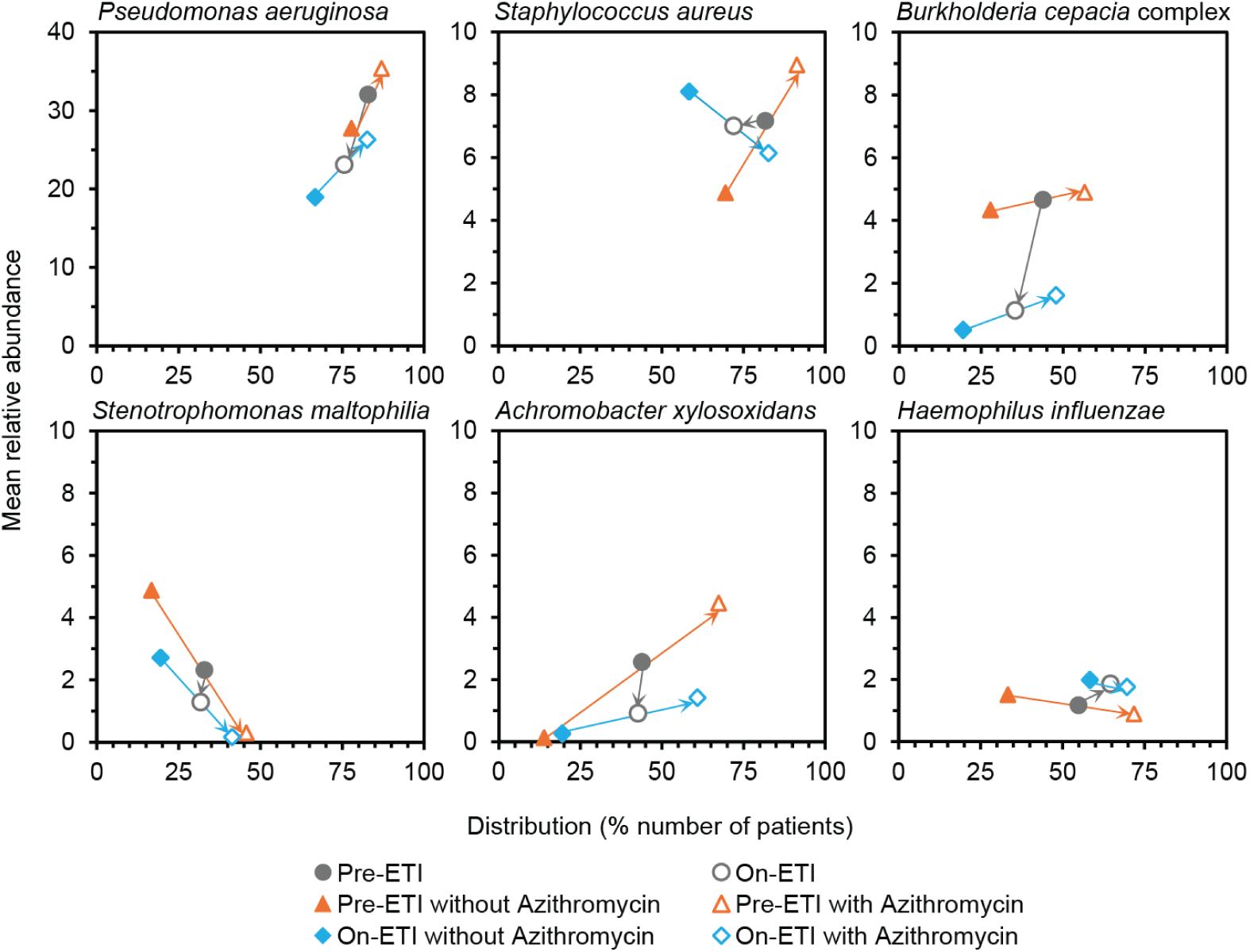
Distribution and abundance of canonical pathogens across adults with CF receiving azithromycin treatment or not when either pre-ETI or on-ETI therapy. In each instance, given is the percentage number of patient respiratory samples each pathogen was observed to be distributed across, plotted against the mean percentage abundance across those samples. In each case, arrows indicate direction of change in distribution and abundance. Distribution-abundance relationships for the whole microbiota for each treatment scenario are given in Supplementary Figure S3 along with the core taxa in each case (Supplementary Table S5).

## Discussion

The introduction of CFTRmt, and particularly the triple drug combination, elexacaftor-tezacaftor-ivacaftor (ETI), have profoundly changed the course of CF for many people living with this inherited disease [21, 37]. Given chronic respiratory infection and concomitant inflammation is the leading cause of morbidity and early mortality for the majority of people living with cystic fibrosis [3–5, 22], there is a pressing need to understand the impact of ETI CFTRmt therapy on the CF airway microbiota. Here for the first time, we assessed the long-term effects (over 3+ years) of ETI therapy on the microbiome ecology of respiratory infection in a large multi-centre and multi-national cohort of awCF incorporating the high interpatient variability inherent in CF [4].

Moving beyond purely comparing pre-ETI versus on-ETI samples we stratified pre-ETI samples by disease severity, a key determinant of microbiome heterogeneity [3–5], and stratified on-ETI samples by duration on therapy. From our previous work, we established there were distinct microbiota characteristics associated with disease severity [4]. Moving from mild to moderate to severe disease CF microbiota exhibited decreasing diversity, increasing dominance, and were progressively dominated by key CF pathogens. With those pathogens becoming ecologically more important in terms of increased abundance and distribution across awCF with increasing severity [4]. Here we found that was the case within the pre-ETI samples (Figure 1 and Table 2), indicating stratification by disease severity was both microbiologically and clinically meaningful for this study. Mean indices of diversity and dominance within on-ETI samples were found to be higher and lower, respectively, than the severe pre-ETI group from 6 months on therapy, and from the mild pre-ETI group from 2 years on therapy (Figure 1). Moreover, CF pathogens typically had progressively reduced ecological importance with longer therapy duration, being replaced by commensal respiratory bacteria, including species of *Prevotella*, *Rothia*, *Streptococcus*, and *Veillonella* genera (Figure 2 and Table 2).

By 3 years no CF pathogens retained core status (except the opportunistic pathogen *H. influenzae*) and all experienced notable reductions in % distribution across patients between 6 months and 3 years (*P. aeruginosa* % distribution from 82% to 36%; *S. aureus* 76% to 31%, *H. influenzae* 66% to 53%, *S. maltophilia* 56% to 14%, *B. cepacia* complex 45% to 10%, and *A. xylosoxidans* 64% to not detected)(Figure 2). While the persistence of all pathogens was reduced and their ecological importance diminished, from an evolutionary perspective a study of *P. aeruginosa* found that strains of this species evolved and new strains emerged with the transition onto ETI therapy [12, 13]. While the clinical significance of this still needs to be elucidated, within a microbiome context it would be reasonable to expect the onset of ETI therapy would place a strong evolutionary selection pressure on not just this one emblematic species but across the majority of species within the CF respiratory microbiota, including other canonical CF pathogens [38]. Despite the notable decreased distribution across awCF, we would recommend future work evaluating whether that selection pressure enables instances of evolution of more virulent strains of respiratory pathogens both typical and non-typical to CF [21].

While with increasing therapy duration on-ETI microbiota characteristics became less like those observed in pre-ETI samples stratified by severity, the on-ETI microbiota characteristics increasingly remodelled towards those observed in the non-CF healthy cohort, though typically remained significantly different (Figure 1 and Figure S2). We posit microbiota remodelling was unlikely to markedly improve in studies with observation timelines of more than three years towards a healthy like state due to factors like irreversible structural lung damage experienced by many awCF with increasing age, the cumulative effects of long-term antibiotic exposure throughout the life of an awCF, as well as current antibiotic treatments all likely contributing to that inhibition [1, 17].

When we examined the individual responses to ETI CFTRmt, lung function and microbiota diversity and dominance generally improved across disease severity groups (Figure 3). With compositional similarity affected by pre-ETI disease severity and long-term therapy duration (Figure 4 and Table 3). Interestingly, antibiotics and lung function, which is indicative of lung damage [1], significantly accounted for variation in respiratory microbiota composition (Table 3). Strikingly, we found that azithromycin therapy specifically exerted a selective pressure on the respiratory microbiota, maintaining a pre-ETI like composition when compared to microbiota compositions without the effect of receiving azithromycin (Figure 4). Also of concern was that this deleterious selective effect also extended to maintaining enhanced CF pathogen persistence regardless of receiving ETI therapy or not (Figure 5). This would suggest that the use of azithromycin when on-ETI therapy both maintains a pre-ETI microbiota dysbiosis and enables elevated persistence of key CF pathogens.

Azithromycin is a macrolide antibiotic that is primarily used in CF, non-CF bronchiectasis, and COPD for its immunomodulatory effects rather than its anti-bacterial properties; with its long-term use associated with reduced frequency of acute pulmonary exacerbations and improved lung function [39]. A recent study investigated the anti-inflammatory effects of azithromycin and ETI, as monotherapy and in combination, and found azithromycin did not provide either antagonistic or synergistic effects when used in combination with ETI [40]. Azithromycin long-term use is associated with toxicities such as hearing loss, the potential for drug-drug interactions, and may even antagonize the anti-Pseudomonal effect of tobramycin [41, 42]. Furthermore, even prior to ETI there was evidence to suggest that the improvements associated with %FEV_1_ and exacerbation frequency of azithromycin may not extend beyond its first year [43]. Subsequently it has been suggested that the CFTRmt era provides an opportunity to re-evaluate therapeutic regimens to identify redundant treatments, and hence reduce polypharmacy burden and safely discontinue treatments without loss of clinical benefit [40]. Here we found that azithromycin had adverse effects on the respiratory microbiota nullifying the observed positive effects of ETI treatment. As such azithromycin appears to be an ideal candidate for discontinuation within the polypharmacy of CF treatment with clinical evaluation required for its continued usage [44, 45].

There are caveats to this study that need to be addressed. First is the use of both sputum and cough swabs within the study. In the pre-CFTRmt era sputum was the preferred common sample type for microbiological analyses, it is now typical that many people with CF receiving ETI are no longer readily sputum productive, even after sputum induction [14, 16, 17]. Moreover, studies that only include individuals that can still produce sputum when on ETI will be biased towards patients with more advanced lung disease and not representative of the wider CF population [14, 17, 46]. To overcome that bias and in the absence of a recommended non-invasive alternative to sputum we utilised a combination of sputum and cough swab samples. As in our previous recent studies, we enhanced DNA recovery from cough swabs by using a modification to the DNA extraction protocol and also assessed the influence of sample type on microbiota analyses [5, 22]. With the exception of sequence reads per sample, we found that microbiota characteristics between the two sample types were not significantly different (Figure S1), as we have previously found [5]. As such we were satisfied both sample types could be used and compared in the current study. However, we would recommend testing the influence of sample type when doing so in each instance.

Another point for consideration is not all samples collected were pre-ETI and on-ETI matched. ETI was first approved either just prior (USA) or during (UK and Canada) the COVID-19 global pandemic, leading to a reduction of patient visits to CF centres and an increase in remote healthcare delivery, and hence disrupted sample acquisition [28, 47]. Additionally, ETI has been widely reported to improve health and extend predicted lifespan for many awCF and as such from our experience many awCF are now less inclined to attend clinic or participate in studies, further adding to the challenge of sample acquisition [8, 48]. Therefore, we took a pragmatic approach deeming any sample acquired would be informative within the ecological analytical framework used for the current study.

Finally, while the bacterial respiratory microbiota has been the central focus for all studies to date, we would recommend that the impacts and consequences of ETI therapy on the poorly understood and understudied fungal microbiota should also be explored. Moreover, as most work to date has focused on adolescents (≥12years) and adults, there is a need to investigate the impact of ETI therapy upon the respiratory microbiota of children with CF (<12 years) who typically have less structural lung damage and do not have long-standing respiratory infection.

## Conclusions

The highly anticipated introduction of the CFTRmt, ETI, has profoundly changed the course of CF for many people living with this inherited disease. What is not certain is the long-term effects of ETI on the respiratory microbiota and the CF pathogens contained within. Across a large multi-centre and multi-national cohort of awCF we determined that the respiratory microbiota became decreasingly like pre-ETI microbiota and remodelled to become more like a healthy microbiota, where canonical CF pathogens progressively become less ecologically important with treatment duration. However, the on-ETI microbiota was impeded from becoming fully healthy like due to continued antibiotic exposure and irreversible lung damage experienced by awCF. Specifically, we discovered that azithromycin had adverse effects on the respiratory microbiota nullifying the observed positive effects of ETI treatment enabling persistence of emblematic CF pathogens. As such azithromycin would be an ideal candidate for discontinuation studies in the CFTRmt era.

## Data Availability

All data produced in the present study are available upon reasonable request to the authors
All data produced in the present work are contained in the manuscript
Anonymised clinical metadata and processed microbiota data have been deposited at https://figshare.com

https://figshare.com/s/bcde4a137d8770f81698.

## Acknowledgements

The authors would like to thank all the patients who took part in this study and the clinical teams at each participating centre. We are grateful to Dr Catherine van der Gast for her constructive and thoughtful comments which have helped to further improve this article.

## Authors’ contributions

CvdG, DWR, and HG conceived the study. CvdG and HG were responsible for microbiota analysis. HG, LRH, MH, GGE, CST, YH, performed sample preparation and analyses. GGE, CST, MDP, JD, JMM, SEL, RWL, AJ, AH, TWVD, and CCT were responsible for sample collection, clinical care records and documentation. HG and CvdG were responsible for the creation of the initial draft of the manuscript. All authors contributed to development of the final manuscript. CvdG is guarantor of this work. The authors read and approved the final manuscript.

## Availability of data and materials

The raw sequence data reported in this study have been deposited in the European Nucleotide Archive under accession numbers PRJEB95094. Anonymised clinical metadata and processed microbiota data have been deposited at https://figshare.com under https://figshare.com/s/bcde4a137d8770f81698.

## Ethics approval and consent to participate

The study was reviewed and approved for each participating centre: Burlington, VT, USA (University of Vermont Institutional Review Board, CHRMS STUDY683), Calgary, AB, Canada (University of Calgary’s Conjoint Health Research Ethics Board, REB-15–2744), Cardiff, UK (Health and Care Research Wales Research Ethics Committee, 18/WA/0089), Manchester, UK (North West – Preston Research Ethics Committee, 20/NW/0302 and NHS Research Ethics Committee North West, Lancaster, 14/NW/1195), Pittsburgh, PA, USA (University of Pittsburgh Institutional Review Board, STUDY19100149), and Southampton, UK (South Central – Hampshire A Research Ethics Committee, 08/H0602/126). All patients attending each participating centre provided written informed consent.

## Consent for publication

Not applicable

## Supplementary Information

**Supplementary Figure 1** Comparisons of microbiota characteristics between cough swab and sputum samples. (A) number of sequence reads between sample types, (B, C, D) comparisons of diversity and dominance, and (E) comparisons of within and between sample type microbiota compositional similarity. Boxplots show 25-75^th^ interquartile (IQR) range with whiskers showing 1.5 times IQR. Purple crosses represent the mean in each group. Circles denote individual measures within a given group. Swab *n* = 102 and sputum *n* = 352. Mean and standard deviation of the mean values: (A) swab = 21632.3 ± 17893.8 and sputum 70333.7 ± 99340.8; (B) swab 8.04 ± 5.81, sputum 9.76 ± 8.60; (C) swab 1.44 ± 0.73, sputum 1.41 ± 0.90; (D) swab 0.44 ± 0.24, sputum 0.48 ± 0.27; and (E) swab 0.26 ± 0.15, sputum 0.26 ± 0.16, between group 0.22 ± 0.14 (Pairwise comparisons: swab = 5151, sputum = 61776, between groups = 35904). Kruskal-Wallis test statistics (*H*) and significance (*P*): (A) *H* = 51.81, *P* <0.0001; (B) *H* = 0.341, *P* =0.559; (C) *H* = 0.133, *P* = 0.715; (D) *H* = 1.115, *P* =0.291; and (E) *H* = 0.271, *P* = 0.603.

**Figure S2** Comparison of microbiota compositional similarities between pre- and on-elexacaftor/tezacaftor/ivacaftor (ETI) sample groups. Respiratory microbiota samples from adults with cystic fibrosis (CF) at (A) 6 months, (B) 1 year, (C) 2 years, and (D) 3 years of therapy duration compared to pre-ETI samples stratified by disease severity, along with samples from a non-CF healthy cohort. Compositional similarities were measured using the Sørensen index of similarity. Boxplots show 25-75^th^ interquartile (IQR) range with whiskers showing 1.5 times IQR. Purple crosses represent the mean in each group. In each instance, circles denote compositional similarities of samples taken pairwise between each given pairing of groups. Summary statistics for PERMANOVA tests between all groups taken pairwise are provided in Supplementary Table S3.

**Figure S3** Distribution and abundance of bacterial taxa across adults with CF receiving Azithromycin treatment or not when either pre-ETI or on-ETI therapy. (**A**) Pre-ETI therapy all samples, (**B**) On-ETI therapy all samples, (**C**) and (**D**) pre-ETI and on-ETI without Azithromycin, and (**E**) and (**F**) pre-ETI and on-ETI with Azithromycin. Given is the percentage number of patient respiratory samples each bacterial taxon was observed to be distributed across, plotted against the mean percentage abundance across those samples. Core taxa are defined as those are in >50% of samples (orange circles), and satellite taxa (grey circles) defined as those that do not. Canonical CF pathogens are highlighted in each plot. Distribution-abundance relationship regression statistics: (**A**) *R*^2^ = 0.62, *F*_1,430_ = 268.4, *P* < 0.0001; (**B**) *R*^2^ = 0.70, *F*_1,377_ = 494.7, *P* < 0.0001; (**C**) *R*^2^ = 0.39, *F*_1,273_ = 171.5, *P* < 0.0001; (**D**) *R*^2^ = 0.53, *F*_1,258_ = 293.4, *P* < 0.0001; (**E**) *R*^2^ = 0.44, *F*_1,374_ = 288.3, *P* < 0.0001; and (**F**) *R*^2^ = 0.61, *F*_1,324_ = 502.2, *P* < 0.0001. Core taxa are listed in Supplementary Table S5.

**Table S1** Kruskal-Wallis summary statistics for all groups taken pairwise. Summary statistics are given for (A) Shannon index of diversity, (B) Simpson’s index of diversity, and (C) Berger-Parker index of dominance (*d*). For each index, test statistic *H* observed is given in the upper triangle and significance (*P*) in the lower triangle. *H* critical was 0.384. Tests with significant differences are highlighted in green.

**Table S2** Core taxa within each on-ETI therapy duration category. (**A**) 6 months duration, (**B**) 1 year, (**C**) 2 years, and (**D**) 3 years duration on ETI therapy. Given is distribution, the percentage number of samples a given core taxon was detected in, and average relative abundance across those samples. Given the length of the ribosomal sequences analysed, species identities should be considered putative. In each instance, taxa are ordered from most to least persistent. Canonical CF pathogens are highlighted in bold.

**Table S3** PERMANOVA summary statistics for all groups taken pairwise. Summary statistics are given for Sørensen indices of similarity between groups *F*-statistic is given in the upper triangle and significance (*P*) in the lower triangle. Tests with significant differences are highlighted in green.

**Table S4** Kruskal-Wallis summary statistics for measures of lung function, diversity and dominance between paired pre- and on-ETI therapy samples when stratified by pre-ETI disease severity status. Summary statistics are given for lung function (%FEV_1_), Shannon index of diversity, Simpson’s index of diversity, and Berger-Parker index of dominance. Given for each test is the Kruskal-Wallis test statistic *H* and significance (*P*). *H* critical was 0.384. Tests with significant differences are highlighted in green.

**Table S5** Core taxa across adults with CF receiving Azithromycin treatment or not when either pre-ETI or on-ETI therapy. Given are core taxa for all pre-ETI and on-ETI therapy samples, pre-ETI therapy not on and on Azithromycin treatment, and on-ETI therapy not on and on Azithromycin treatment. Distribution (Dis) is the percentage number of samples a given core taxon was detected in, and average relative abundance (Abu) across those samples. Given the length of the ribosomal sequences analysed, species identities should be considered putative. In each instance, taxa are ordered by distribution (from most to least). Canonical CF pathogens are highlighted in bold.

