## Supplemental Materials for "Remodelling of cystic fibrosis respiratory microbiota in response to extended Elexacaftor–Tezacaftor–Ivacaftor therapy"

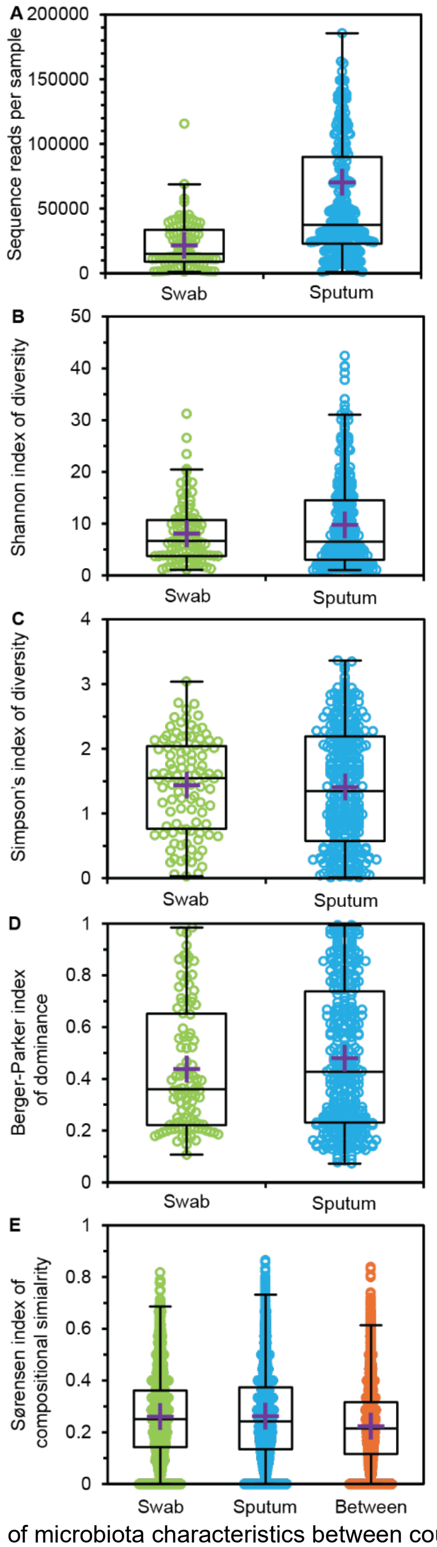


**Supplementary Figure 1** Comparisons of microbiota characteristics between cough swab and sputum samples. (A) number of sequence reads between sample types, (B, C, D) comparisons of diversity and dominance, and (E) comparisons of within and between sample type microbiota compositional similarity. Boxplots show 25-75$^{th}$ interquartile (IQR) range with whiskers showing 1.5 times IQR. Purple crosses represent the mean in each group. Circles denote individual measures within a given group. Swab $n$ = 102 and sputum $n$ = 352. Mean and standard deviation of the mean values: (A) swab = 21632.3 ± 17893.8 and sputum 70333.7 ± 99340.8; (B) swab 8.04 ± 5.81, sputum 9.76 ± 8.60; (C) swab 1.44 ± 0.73, sputum 1.41 ± 0.90; (D) swab 0.44 ± 0.24, sputum 0.48 ± 0.27; and (E) swab 0.26 ± 0.15, sputum 0.26 ± 0.16, between group 0.22 ± 0.14 (Pairwise comparisons: swab = 5151, sputum = 61776, between groups = 35904). Kruskal-Wallis test statistics ($H$) and significance ($P$): (A) $H$ = 51.81, $P$ <0.0001; (B) $H$ = 0.341, $P$ =0.559; (C) $H$ = 0.133, $P$ = 0.715; (D) $H$ = 1.115, $P$ =0.291; and (E) $H$ = 0.271, $P$ = 0.603.

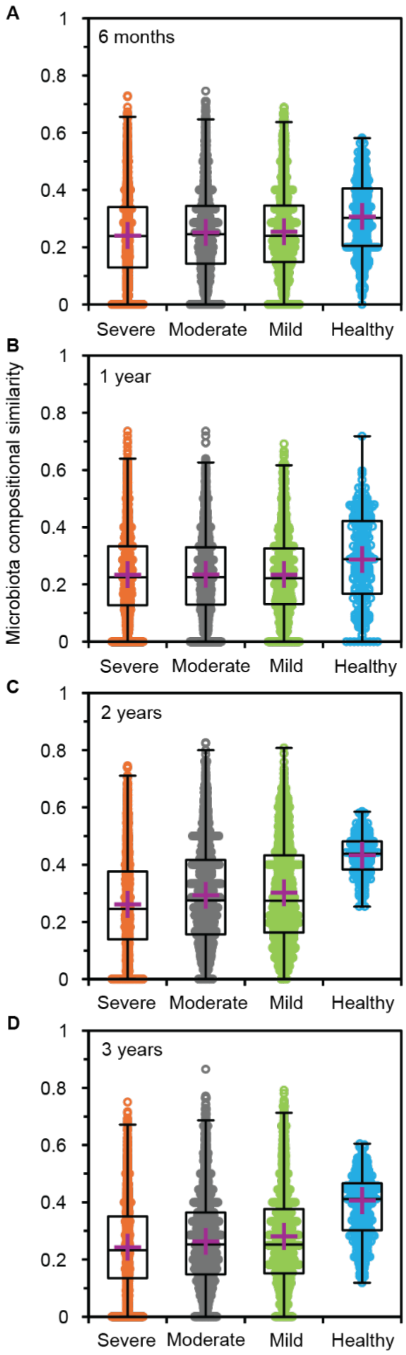


**Figure S2** Comparison of microbiota compositional similarities between pre- and on-elexacaftor/tezacaftor/ivacaftor (ETI) sample groups. Respiratory microbiota samples from adults with cystic fibrosis (CF) at (A) 6 months, (B) 1 year, (C) 2 years, and (D) 3 years of therapy duration compared to pre-ETI samples stratified by disease severity, along with samples from a non-CF healthy cohort. Compositional similarities were measured using the Sørensen index of similarity. Boxplots show 25-75$^{th}$ interquartile (IQR) range with whiskers showing 1.5 times IQR. Purple crosses represent the mean in each group. In each instance, circles denote compositional similarities of samples taken pairwise between each given pairing of groups. Summary statistics for PERMANOVA tests between all groups taken pairwise are provided in Supplementary Table S3.

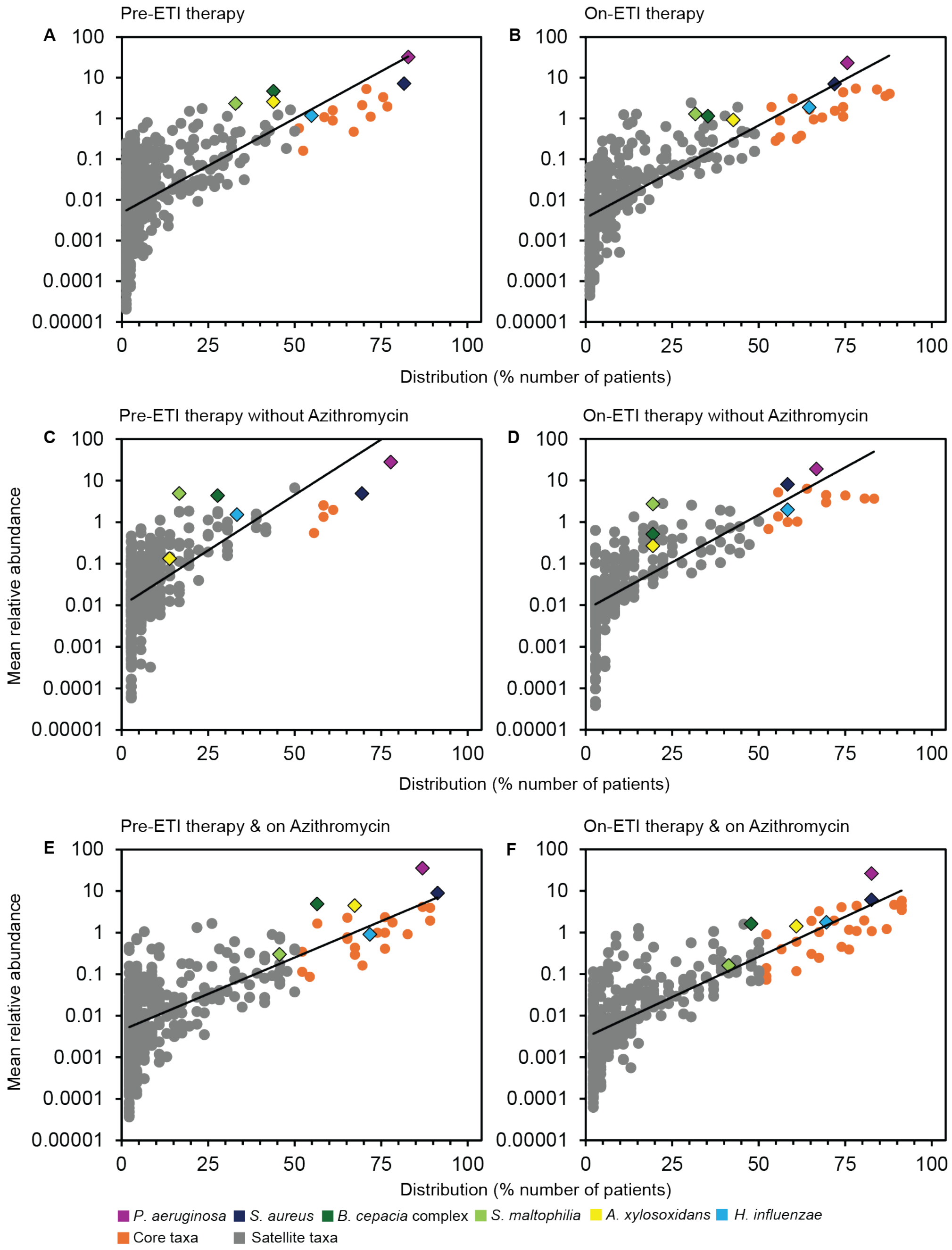


**Figure S3** Distribution and abundance of bacterial taxa across adults with CF receiving Azithromycin treatment or not when either pre-ETI or on-ETI therapy. (**A**) Pre-ETI therapy all samples, (**B**) On-ETI therapy all samples, (**C**) and (**D**) pre-ETI and on-ETI without Azithromycin, and (**E**) and (**F**) pre-ETI and on-ETI with Azithromycin. Given is the percentage number of patient respiratory samples each bacterial taxon was observed to be distributed across, plotted against the mean percentage abundance across those samples. Core taxa are defined as those are in >50% of samples (orange circles), and satellite taxa (grey circles) defined as those that do not. Canonical CF pathogens are highlighted in each plot. Distribution-abundance relationship regression statistics: (**A**) $R^2$ = 0.62, $F_{1,430}$ = 268.4, $P < 0.0001$; (**B**) $R^2$ = 0.70, $F_{1,377}$ = 494.7, $P < 0.0001$; (**C**) $R^2$ = 0.39, $F_{1,273}$ = 171.5, $P < 0.0001$; (**D**) $R^2$ = 0.53, $F_{1,258}$ = 293.4, $P < 0.0001$; (**E**) $R^2$ = 0.44, $F_{1,374}$ = 288.3, $P < 0.0001$; and (**F**) $R^2$ = 0.61, $F_{1,324}$ = 502.2, $P < 0.0001$. Core taxa are listed in Supplementary Table S5.

**Table S1** Kruskal-Wallis summary statistics for all groups taken pairwise. Summary statistics are given for (A) Shannon index of diversity, (B) Simpson's index of diversity, and (C) Berger-Parker index of dominance (*d*). For each index, test statistic *H* observed is given in the upper triangle and significance (*P*) in the lower triangle. Tests with significant differences are highlighted in green.

| **A** | | **Pre-ETI** | | | **Non-CF** | **On-ETI** | | | |
|---|---|---|---|---|---|---|---|---|---|
| | | **Severe** | **Moderate** | **Mild** | **Healthy** | **6M** | **1Y** | **2Y** | **3Y** |
| **Pre-ETI** | **Severe** | | 0.072 | 3.991 | 18.904 | 0.796 | 1.461 | 22.629 | 9.928 |
| | **Moderate** | 0.789 | | 4.307 | 19.292 | 0.883 | 2.690 | 26.518 | 12.553 |
| | **Mild** | 0.045 | 0.038 | | 17.128 | 0.067 | 0.001 | 10.862 | 5.328 |
| **Non-CF** | **Healthy** | <0.0001 | <0.0001 | <0.0001 | | 13.994 | 13.092 | 7.400 | 7.552 |
| **On-ETI** | **6M** | 0.372 | 0.267 | 0.795 | <0.0001 | | 0.229 | 7.775 | 4.629 |
| | **1Y** | 0.227 | 0.101 | 0.973 | <0.0001 | 0.585 | | 7.890 | 3.912 |
| | **2Y** | <0.0001 | <0.0001 | <0.0001 | 0.007 | 0.005 | 0.005 | | 0.452 |
| | **3Y** | 0.002 | <0.0001 | 0.021 | 0.006 | 0.031 | 0.046 | 0.501 | |

| **B**<br>**Simpson's index** | | **Pre-ETI** | | | **Non-CF** | **On-ETI** | | | |
|---|---|---|---|---|---|---|---|---|---|
| | | **Severe** | **Moderate** | **Mild** | **Healthy** | **6M** | **1Y** | **2Y** | **3Y** |
| **Pre-ETI** | **Severe** | | 0.096 | 3.968 | 15.597 | 0.379 | 1.535 | 17.975 | 9.193 |
| | **Moderate** | 0.756 | | 5.024 | 16.824 | 0.883 | 2.593 | 21.768 | 11.252 |
| | **Mild** | 0.047 | 0.025 | | 13.298 | 0.407 | 0.001 | 14.733 | 4.353 |
| **Non-CF** | **Healthy** | <0.0001 | <0.0001 | <0.0001 | | 12.522 | 10.982 | 3.717 | 4.343 |
| **On-ETI** | **6M** | 0.539 | 0.347 | 0.524 | <0.0001 | | 0.536 | 11.67 | 5.465 |
| | **1Y** | 0.215 | 0.107 | 0.992 | <0.0001 | 0.464 | | 9.249 | 3.923 |
| | **2Y** | <0.0001 | <0.0001 | 0.001 | 0.054 | 0.001 | 0.002 | | 1.073 |
| | **3Y** | 0.002 | 0.001 | 0.037 | 0.037 | 0.002 | 0.048 | 0.300 | |

| **C**<br>**Berger-Parker** | | **Pre-ETI** | | | **Non-CF** | **On-ETI** | | | |
|---|---|---|---|---|---|---|---|---|---|
| | | **Severe** | **Moderate** | **Mild** | **Healthy** | **6M** | **1Y** | **2Y** | **3Y** |
| **Pre-ETI** | **Severe** | | 0.171 | 2.526 | 12.667 | 0.348 | 1.205 | 21.244 | 8.700 |
| | **Moderate** | 3.841 | | 5.995 | 14.183 | 1.286 | 2.806 | 27.959 | 11.615 |
| | **Mild** | 0.112 | 0.014 | | 10.214 | 0.384 | 0.023 | 17.165 | 4.135 |
| **Non-CF** | **Healthy** | <0.0001 | <0.0001 | <0.0001 | | 8.836 | 9.056 | 0.628 | 2.795 |
| **On-ETI** | **6M** | 0.555 | 0.257 | 0.535 | 0.003 | | 0.329 | 12.379 | 4.990 |
| | **1Y** | 0.272 | 0.094 | 0.879 | 0.003 | 0.566 | | 11.601 | 3.901 |
| | **2Y** | <0.0001 | <0.0001 | <0.0001 | 0.428 | <0.0001 | 0.001 | | 1.868 |
| | **3Y** | 0.003 | 0.001 | 0.042 | 0.095 | 0.025 | 0.048 | 0.172 | |

**Table S2** Core taxa within each on-ETI therapy duration category**.** (**A**) 6 months duration, (**B**) 1 year, (**C**) 2 years, and (**D**) 3 years duration on ETI therapy. Given is distribution, the percentage number of samples a given core taxon was detected in, and average relative abundance across those samples. Given the length of the ribosomal sequences analysed, species identities should be considered putative. In each instance, taxa are ordered from most to least persistent. Canonical CF pathogens are highlighted in bold.

**(A)** 6 months

| Taxon | Distribution | Abundance |
|---|---|---|
| *Rothia mucilaginosa* | 93.5 | 3.7 |
| ***Pseudomonas aeruginosa*** | 87.0 | 30.5 |
| *Veillonella dispar* | 84.8 | 5.5 |
| *Streptococcus salivarius* | 84.8 | 4.8 |
| *Streptococcus parasanguinis* | 82.6 | 4.4 |
| *Veillonella parvula* | 82.6 | 2.1 |
| *Rothia dentocariosa* | 82.6 | 1.2 |
| *Prevotella salivae* | 80.4 | 1.2 |
| *Prevotella melaninogenica* | 76.1 | 4.0 |
| ***Staphylococcus aureus*** | 76.1 | 3.8 |
| *Gemella parahaemolysans* | 76.1 | 1.4 |
| *Granulicatella adiacens* | 73.9 | 0.9 |
| *Lancefieldella parvula* | 71.7 | 0.1 |
| *Oribacterium sinus* | 69.6 | 0.2 |
| *Prevotella oris* | 67.4 | 1.2 |
| *Prevotella pallens* | 67.4 | 1.1 |
| ***Haemophilus influenzae*** | 67.4 | 1.0 |
| *Actinomyces naeslundii* | 67.4 | 0.4 |
| *Prevotella histicola* | 65.2 | 3.7 |
| *Streptococcus peroris* | 65.2 | 2.7 |
| *Schaalia odontolytica* | 65.2 | 0.7 |
| ***Achromobacter xylosoxidans*** | 63.0 | 1.5 |
| *Scardovia wiggsiae* | 63.0 | 0.2 |
| ***Burkholderia cepacia* complex** | 56.5 | 2.0 |
| *Fusobacterium nucleatum* | 56.5 | 0.6 |
| *Anaeroglobus micronuciformis* | 56.5 | 0.5 |
| *Prevotella denticola* | 56.5 | 0.4 |
| *Campylobacter concisus* | 56.5 | 0.4 |
| *Neisseria cinerea* | 54.3 | 0.9 |
| *Lautropia mirabilis* | 54.3 | 0.4 |
| *Solobacterium moorei* | 54.3 | 0.1 |
| *Actinomyces graevenitzii* | 52.2 | 0.3 |
| *Lachnoanaerobaculum orale* | 52.2 | 0.1 |

**(B)** 1 year

| Taxon | Distribution | Abundance |
|---|---|---|
| *Veillonella dispar* | 89.7 | 6.3 |
| *Streptococcus salivarius* | 87.2 | 11.7 |
| *Rothia mucilaginosa* | 82.1 | 2.9 |
| *Streptococcus parasanguinis* | 74.4 | 5.8 |
| *Prevotella melaninogenica* | 74.4 | 4.5 |
| *Gemella parahaemolysans* | 71.8 | 2.0 |
| *Schaalia odontolytica* | 71.8 | 1.7 |
| *Veillonella parvula* | 61.5 | 1.9 |
| *Rothia dentocariosa* | 61.5 | 1.1 |
| *Granulicatella adiacens* | 59.0 | 0.6 |
| *Lancefieldella parvula* | 59.0 | 0.3 |
| ***Pseudomonas aeruginosa*** | 56.4 | 17.8 |
| *Prevotella histicola* | 53.8 | 2.2 |
| ***Haemophilus influenzae*** | 53.8 | 1.5 |
| *Prevotella pallens* | 53.8 | 0.5 |
| ***Staphylococcus aureus*** | 51.3 | 6.3 |

**(C)** 2 years

| Taxon | Distribution | Abundance |
|---|---|---|
| *Streptococcus parasanguinis* | 87.5 | 3.2 |
| *Streptococcus oralis* | 85.0 | 8.7 |
| *Veillonella dispar* | 85.0 | 4.7 |
| *Granulicatella adiacens* | 85.0 | 3.1 |
| ***Pseudomonas aeruginosa*** | 80.0 | 12.0 |
| *Rothia mucilaginosa* | 80.0 | 2.1 |
| *Streptococcus salivarius* | 72.5 | 2.9 |
| *Gemella parahaemolysans* | 72.5 | 1.5 |
| ***Staphylococcus aureus*** | 67.5 | 12.2 |
| ***Haemophilus influenzae*** | 65.0 | 2.3 |
| *Schaalia odontolytica* | 65.0 | 1.1 |
| *Rothia dentocariosa* | 60.0 | 0.8 |
| *Prevotella melaninogenica* | 57.5 | 4.0 |
| *Veillonella parvula* | 55.0 | 1.7 |
| *Prevotella salivae* | 52.5 | 0.9 |

**(D)** 3 years

| Taxon | Distribution | Abundance |
|---|---|---|
| *Streptococcus parasanguinis* | 100.0 | 7.5 |
| *Rothia mucilaginosa* | 97.4 | 6.9 |
| *Granulicatella adiacens* | 94.7 | 3.7 |
| *Veillonella dispar* | 92.1 | 7.5 |
| *Streptococcus salivarius* | 81.6 | 9.4 |
| *Prevotella melaninogenica* | 81.6 | 5.0 |
| *Schaalia odontolytica* | 76.3 | 1.8 |
| *Gemella parahaemolysans* | 68.4 | 2.1 |
| *Prevotella histicola* | 63.2 | 3.1 |
| *Campylobacter concisus* | 60.5 | 0.7 |
| *Stomatobaculum longum* | 60.5 | 0.6 |
| *Rothia dentocariosa* | 57.9 | 1.5 |
| *Prevotella salivae* | 57.9 | 0.8 |
| *Lancefieldella parvula* | 57.9 | 0.7 |
| *Veillonella parvula* | 55.3 | 1.2 |
| *Oribacterium sinus* | 55.3 | 0.6 |
| *Streptococcus oralis* | 52.6 | 3.4 |
| ***Haemophilus influenzae*** | 52.6 | 3.1 |
| *Lachnoanaerobaculum orale* | 52.6 | 0.9 |

**Table S3** PERMANOVA summary statistics for all groups taken pairwise. Summary statistics are given for Sørensen indices of similarity between groups *F*-statistic is given in the upper triangle and significance (*P*) in the lower triangle. Tests with significant differences are highlighted in green.

| | | Pre-ETI | | | Non-CF | On-ETI | | | |
|---|---|---|---|---|---|---|---|---|---|
| | | **Severe** | **Moderate** | **Mild** | **Healthy** | **6M** | **1Y** | **2Y** | **3Y** |
| **Pre-ETI** | **Severe** | | 1.51 | 2.90 | 8.25 | 8.11 | 4.39 | 6.15 | 4.95 |
| | **Moderate** | 0.0877 | | 1.84 | 7.59 | 6.14 | 3.62 | 6.51 | 5.06 |
| | **Mild** | 0.0011 | 0.0327 | | 5.99 | 4.07 | 1.82 | 3.83 | 3.94 |
| **Non-CF** | **Healthy** | <0.0001 | <0.0001 | <0.0001 | | 6.49 | 5.89 | 6.84 | 8.31 |
| **On-ETI** | **6M** | <0.0001 | <0.0001 | 0.0002 | <0.0001 | | 2.64 | 6.14 | 6.83 |
| | **1Y** | <0.0001 | 0.0003 | 0.0249 | <0.0001 | 0.0038 | | 1.87 | 3.44 |
| | **2Y** | <0.0001 | <0.0001 | <0.0001 | <0.0001 | <0.0001 | 0.0302 | | 2.76 |
| | **3Y** | <0.0001 | <0.0001 | 0.0002 | <0.0001 | <0.0001 | <0.0001 | 0.0018 | |

**Table S4** Kruskal-Wallis summary statistics for measures of lung function, diversity and dominance between paired pre- and on-ETI therapy samples when stratified by pre-ETI disease severity status. Summary statistics are given for lung function (%$FEV_1$), Shannon index of diversity, Simpson's index of diversity, and Berger-Parker index of dominance. Given for each test is the Kruskal-Wallis test statistic *H* and significance (*P*). Tests with significant differences are highlighted in green.

| | Severe | | Moderate | | Mild | |
|---|---|---|---|---|---|---|
| | *H* | *P* | *H* | *P* | *H* | *P* |
| %$FEV_1$ | 16.276 | < 0.0001 | 10.484 | 0.001 | 4.996 | 0.025 |
| Shannon index | 0.115 | 0.734 | 0.798 | 0.372 | 5.523 | 0.019 |
| Simpson's index | 0.158 | 0.691 | 0.744 | 0.389 | 4.731 | 0.030 |
| Berger-Parker index | 0.034 | 0.854 | 6.088 | 0.014 | 0.793 | 0.373 |

**Table S5** Core taxa across adults with CF receiving Azithromycin treatment or not when either pre-ETI or on-ETI therapy. Given are core taxa for all pre-ETI and on-ETI therapy samples, pre-ETI therapy not on and on Azithromycin treatment, and on-ETI therapy not on and on Azithromycin treatment. Distribution (Dis) is the percentage number of samples a given core taxon was detected in, and average relative abundance (Abu) across those samples. Given the length of the ribosomal sequences analysed, species identities should be considered putative. In each instance, taxa are ordered by distribution (from most to least). Canonical CF pathogens are highlighted in bold.

| Pre-ETI (All samples) | Dis | Abu | On-ETI (All samples) | Dis | Abu | Pre-ETI without Azithromycin | Dis | Abu | Pre-ETI with Azithromycin | Dis | Abu | On-ETI without Azithromycin | Dis | Abu | On-ETI with Azithromycin | Dis | Abu |
|---|---|---|---|---|---|---|---|---|---|---|---|---|---|---|---|---|---|
| ***Pseudomonas aeruginosa*** | 82.9 | 32.0 | *Streptococcus parasanguinis* | 87.8 | 4.1 | ***Pseudomonas aeruginosa*** | 77.8 | 27.8 | ***Staphylococcus aureus*** | 91.3 | 9.0 | *Streptococcus parasanguinis* | 83.3 | 3.7 | *Veillonella dispar* | 91.3 | 5.8 |
| ***Staphylococcus aureus*** | 81.7 | 7.2 | *Rothia mucilaginosa* | 86.6 | 3.6 | ***Staphylococcus aureus*** | 69.4 | 4.9 | *Veillonella dispar* | 89.1 | 4.0 | *Rothia mucilaginosa* | 80.6 | 3.7 | *Streptococcus parasanguinis* | 91.3 | 4.4 |
| *Rothia mucilaginosa* | 76.8 | 2.0 | *Veillonella dispar* | 84.1 | 5.2 | *Rothia mucilaginosa* | 61.1 | 2.0 | *Rothia mucilaginosa* | 89.1 | 2.0 | *Veillonella dispar* | 75.0 | 4.3 | *Rothia mucilaginosa* | 91.3 | 3.5 |
| *Veillonella dispar* | 75.6 | 3.3 | *Streptococcus salivarius* | 78.0 | 5.4 | *Streptococcus parasanguinis* | 58.3 | 2.6 | ***Pseudomonas aeruginosa*** | 87.0 | 35.4 | *Prevotella melaninogenica* | 69.4 | 4.5 | *Streptococcus salivarius* | 89.1 | 4.7 |
| *Granulicatella adiacens* | 72.0 | 1.1 | ***Pseudomonas aeruginosa*** | 75.6 | 23.1 | *Veillonella dispar* | 58.3 | 2.5 | *Streptococcus salivarius* | 87.0 | 4.2 | *Granulicatella adiacens* | 69.4 | 3.0 | *Rothia dentocariosa* | 87.0 | 1.2 |
| *Streptococcus salivarius* | 70.7 | 5.3 | *Prevotella melaninogenica* | 74.4 | 4.4 | *Granulicatella adiacens* | 58.3 | 1.4 | *Granulicatella adiacens* | 82.6 | 0.9 | ***Pseudomonas aeruginosa*** | 66.7 | 18.9 | ***Pseudomonas aeruginosa*** | 82.6 | 26.3 |
| *Streptococcus parasanguinis* | 69.5 | 2.1 | *Granulicatella adiacens* | 74.4 | 1.9 | *Rothia dentocariosa* | 55.6 | 0.6 | *Streptococcus parasanguinis* | 78.3 | 1.8 | *Streptococcus salivarius* | 63.9 | 6.4 | ***Staphylococcus aureus*** | 82.6 | 6.1 |
| *Rothia dentocariosa* | 67.1 | 0.5 | *Rothia dentocariosa* | 74.4 | 1.1 | | | | *Prevotella melaninogenica* | 76.1 | 2.4 | *Gemella parahaemolysans* | 61.1 | 1.0 | *Prevotella salivae* | 82.6 | 1.1 |
| *Prevotella melaninogenica* | 61.0 | 1.6 | ***Staphylococcus aureus*** | 72.0 | 7.0 | | | | *Gemella parahaemolysans* | 76.1 | 1.0 | ***Staphylococcus aureus*** | 58.3 | 8.1 | *Gemella parahaemolysans* | 80.4 | 2.0 |
| *Gemella parahaemolysans* | 61.0 | 0.9 | *Gemella parahaemolysans* | 72.0 | 1.5 | | | | *Rothia dentocariosa* | 76.1 | 0.4 | ***Haemophilus influenzae*** | 58.3 | 2.0 | *Prevotella melaninogenica* | 78.3 | 4.4 |
| *Veillonella parvula* | 58.5 | 1.1 | *Prevotella salivae* | 68.3 | 1.1 | | | | *Veillonella parvula* | 73.9 | 1.0 | *Rothia dentocariosa* | 58.3 | 1.0 | *Granulicatella adiacens* | 78.3 | 1.1 |
| ***Haemophilus influenzae*** | 54.9 | 1.2 | *Schaalia odontolytica* | 65.9 | 1.0 | | | | ***Haemophilus influenzae*** | 71.7 | 0.9 | *Streptococcus oralis* | 55.6 | 5.2 | *Schaalia odontolytica* | 76.1 | 1.2 |
| *Campylobacter concisus* | 52.4 | 0.2 | ***Haemophilus influenzae*** | 64.6 | 1.9 | | | | *Campylobacter concisus* | 69.6 | 0.2 | *Veillonella parvula* | 55.6 | 1.4 | *Oribacterium sinus* | 76.1 | 0.4 |
| *Prevotella salivae* | 51.2 | 0.6 | *Veillonella parvula* | 64.6 | 1.7 | | | | ***Achromobacter xylosoxidans*** | 67.4 | 4.5 | *Schaalia odontolytica* | 52.8 | 0.7 | *Prevotella histicola* | 73.9 | 4.1 |
| | | | *Lancefieldella parvula* | 62.2 | 0.4 | | | | *Prevotella salivae* | 67.4 | 0.4 | | | | *Lancefieldella parvula* | 73.9 | 0.5 |
| | | | *Oribacterium sinus* | 61.0 | 0.3 | | | | *Oribacterium sinus* | 67.4 | 0.3 | | | | *Veillonella parvula* | 71.7 | 1.9 |
| | | | *Prevotella histicola* | 59.8 | 3.1 | | | | *Prevotella oris* | 65.2 | 2.3 | | | | ***Haemophilus influenzae*** | 69.6 | 1.8 |
| | | | *Prevotella pallens* | 56.1 | 0.9 | | | | *Streptococcus peroris* | 65.2 | 0.8 | | | | *Streptococcus peroris* | 67.4 | 3.3 |
| | | | *Campylobacter concisus* | 56.1 | 0.3 | | | | *Schaalia odontolytica* | 65.2 | 0.7 | | | | *Prevotella pallens* | 67.4 | 1.0 |
| | | | *Actinomyces naeslundii* | 54.9 | 0.3 | | | | ***Burkholderia cepacia complex*** | 56.5 | 4.9 | | | | *Actinomyces naeslundii* | 67.4 | 0.2 |
| | | | *Prevotella oris* | 53.7 | 1.9 | | | | *Prevotella histicola* | 56.5 | 1.7 | | | | *Prevotella oris* | 65.2 | 1.9 |
| | | | | | | | | | *Lancefieldella parvula* | 54.3 | 0.1 | | | | *Campylobacter concisus* | 65.2 | 0.3 |
| | | | | | | | | | *Prevotella pallens* | 52.2 | 0.3 | | | | ***Achromobacter xylosoxidans*** | 60.9 | 1.4 |
| | | | | | | | | | *Prevotella denticola* | 52.2 | 0.1 | | | | *Prevotella denticola* | 60.9 | 0.6 |
| | | | | | | | | | | | | | | | *Capnocytophaga gingivalis* | 60.9 | 0.1 |
| | | | | | | | | | | | | | | | *Anaeroglobus micronuciformis* | 56.5 | 0.4 |
| | | | | | | | | | | | | | | | *Neisseria cinerea* | 52.2 | 0.9 |
| | | | | | | | | | | | | | | | *Stomatobaculum longum* | 52.2 | 0.1 |
| | | | | | | | | | | | | | | | *Solobacterium moorei* | 52.2 | 0.1 |
| | | | | | | | | | | | | | | | *Lachnoanaerobaculum gingivalis* | 52.2 | 0.1 |
